# Comparative efficacy of interferential therapy, bronchodilators, and body positioning on asthma control and quality of life of patients with bronchial asthma: Study protocol

**DOI:** 10.1101/2021.11.29.21267003

**Authors:** Eniola Awolola Oladejo, Sonill Maharaj Sooknunan

## Abstract

**Background:** Interferential therapy (IFT) is the application of two medium frequency currents to the skin to stimulate and activate different systems in the body using specific frequencies and frequency ranges. IFT aims to reduce myalgia in the chest and upper back, reduce muscular fatigue and induce mucus expectoration. This study is designed to compare the efficacy of IFT on asthma control and quality of life of asthma patients pre-and post-administration of bronchodilators and determine the most influential body position in its application.

**Methods:** Forty-eight (48) patients aged 18 years and above with bronchial Asthma attending the respiratory clinic at the Lagos State University Teaching Hospital, Ikeja (LASUTH) will be assessed for the study eligibility. The study design will be a double-blinded, randomized control trial with four intervention groups and four parallel placebo control groups. IFT will be administered as an intervention to patients on short and long-acting bronchodilators in an assigned fundamental body position for 20 minutes. Six continuous outcome variables at different points will be utilized as an outcome measure. Baseline Pulmonary Function Test (PFT) will be assessed on entry into the study, Quality of life and asthma control will be evaluated every two (2) weeks of the study. Data obtained will be analyzed using descriptive and inferential statistics of repeated ANOVA; P<0.05.

**Discussion:** The study outcome will compare the efficacy of IFT on Bronchial Asthma, identify its effect in different body positions, and compare the relationship between its application and the bronchodilator medication frequently used by the patients.

**Trial Registration:** **https://pactr.samrc.ac.za/TrialDisplay.aspx?TrialID=10942:**

PACTR202005807526130

## Background

Worldwide, approximately 300 million people are affected with bronchial Asthma ^1^; It has been more prevalent in developed countries, with higher rates seen in Australia, UK, and New Zealand ^1^. In the Nigerian population, the prevalence of Asthma ranges from 7% to 18% ^1-4^.

According to Alexander.O. Oni1 ^5^, Asthma affects all age groups, races, and sex; for unknown reasons, boys are usually more affected than girls. The prevalence becomes equal by the third decade of life; more women than men are affected after that.

Symptom relief, reduction in the use of on-demand inhalers, improvement in activities, and lung function is the day-to-day Asthma control achievement ^6^. The absence of asthma exacerbations ensures the minimization of future risks by preventing the accelerated decline in lung function and the side-effects from medications over time ^6^.

Progress is being made in the understanding and management of Asthma, the inflammatory nature of the disease, use of steroids, and add-on of inhaled bronchodilator combined with steroids, devices to deliver the medications appropriately, and appreciation of the value of self-management education ^1, 5^.

In the treatment of airways disorders, bronchodilators are central; they are the mainstay of managing most chronic obstructive pulmonary disease and are critical in managing Asthma. Bronchodilators have a direct relaxation effect on airway smooth muscle cells. β (2)-adrenoceptor (AR) agonists, muscarinic receptor antagonists, and xanthines which can be used individually or in combination are three major classes of bronchodilators. The inhaled route of administration is currently considered to minimize systemic effects. For immediate rescue of symptoms, fast- and short-acting agents are preferred, while long-acting agents are best used for maintenance therapy ^7^.

According to Boros and Martusewicz-Boros ^8^, airway reversibility is a test commonly used in diagnosing obstructive lung disease; its result can be used as a differential diagnosis between Asthma and chronic obstructive pulmonary disease. Chung, Wenzel ^9^ identified FEV1/FVC<LLN or 0.70 or <LLN as a tool for diagnosing airway obstruction. Significant improvement (positive bronchodilatation test) is defined as an increase in the FEV1 or FVC of more than 200 mL, and 12% predicted ^9^; however, another method of assessment (200 mL and 12% of baseline) are also mentioned and may be used ^10, 11^.

Several studies conducted to understand the effect of body position on pulmonary function, the most recent being a systematic review by Katz, Arish ^12^, who reported a higher FEV1, FVC, FRC, Imax, and PEF values in most studies involving healthy subjects or patients with lung, heart, neuromuscular disease, or obesity in a more erect position. For subjects with tetraplegia, spinal cord injury, FVC, and FEV1 were higher in supine vs. sitting ^12^.

Interferential therapy (IFT) involves the application of two medium frequency currents to the skin in such a way that the currents “interfere” with each other to produce a “beat” frequency ^13^. The difference between the medium frequency currents is termed the beat frequency, and the body recognizes it as the required low-frequency current. Interferential therapy induces expectoration by making sputum on the bronchi surface mobile, it reduces shoulder stiffness, muscular fatigue, and myalgia in the chest and upper back regions ^14^.

Interferential current (IFC), a non-invasive treatment modality often used to induce analgesia, elicit muscle contractions, and reduce edema ^15-17^. For many years, asthma medications’ effectiveness has been assessed by measuring their impact on expected clinical outcomes such as expiratory flow rates, symptoms, the need for other medications, and airway responsiveness ^18^. This impact is very important, but none of them tells us whether the patients can function better in their day-to-day life ^18^.

This study is designed to investigate the efficacy of a non-invasive therapeutic modality on airway reversibility, asthma control, quality of life of bronchial asthma patients in 2 different body positions using the GLI (LLN) 2012 reference equation for Asthma classification and the 2019 Pan African Thoracic Society (PATS) and European Respiratory society guideline for the validation of spirometry test results. This study’s outcome may provide a non-invasive solution to the bronchospasm frequently experienced during an asthma attack.

### Statement of the problem

In managing Asthma, healthcare providers and patients encounter challenges, including challenges in diagnosis, treatment, follow-up, and other general challenges ^19^. Interferential Therapy (IFT) is a valuable treatment system for many years ^13^. Over the years, research has revealed that Asthma’s quality of life is a very distinct component of overall asthma status ^18^. In a study by Aweto, Tella ^20^ on the effect of interferential therapy on selected cardiopulmonary parameters, asthma control, and quality of life of people living with bronchial Asthma for six weeks, a statistical improvement was recorded in asthma control, quality of life and some of the pulmonary parameters of the patients. It will be necessary to identify the possible interaction of IFT with or without BD frequently used by asthma patients, perform a post-BD and IFT airway reversibility challenge test and evaluate the effect of IFT on varying key body positions, and conduct the study over an extended duration of time to improve the reliability of the study. Another study by Mohammed and Elyazed ^21^, at the Abu El Reish pediatric hospital compared the efficacy of LASER puncture therapy versus Interferential therapy as a combined treatment in Asthmatic Egyptian children revealed a significant improvement in both treatment approaches over diaphragmatic exercises, however, the study did not consider the participants’ medication’s possible effect during the intervention. Both studies did not consider the criterion required for an acceptable, useable and repeatable Pulmonary function Test. The instruments used for the Pulmonary function test did not indicate the record of daily equipment calibrations as recommended by ATS/ERS 2005 guideline for a valid device volume and linearity update. The above factors may affect the test’s reliability, and the results may not be a true reflection of the participants’ ventilatory performance.

This study is therefore designed to investigate the efficacy of IFT on Airway reversibility, Asthma control, quality of life of Bronchial Asthma patients who receive Interferential Therapy for three months in two (2) different body positions using the GLI (LLN) 2012 reference equation for Asthma classification and the 2019 Pan African Thoracic Society (PATS) and European Respiratory society guideline for the validation of spirometry test results ^22, 23^. This study’s outcome may provide a non-invasive solution to the symptoms frequently experienced during an asthma attack.

### Aim of the study

The overall aim of the study is to determine the effect of IFT applied in 45° or 90° long sitting with a bronchodilator (short or long acting) on Asthma control, quality of life, and selected pulmonary variables of Asthmatic patients attending the respiratory clinic of Lagos State University Teaching hospital, Ikeja. Lagos.

### Hypotheses

#### Ho

Interferential therapy (IFT) applied in 45°, or 90° long sitting will have no significant effect on airway reversibility, asthma control, and bronchial asthma patients’ quality of life.

#### Ha

Interferential therapy (IFT) applied in 45°, or 90° long sitting will have no significant effect on airway reversibility, asthma control, and bronchial asthma patients’ quality of life.

#### Delimitation

This study will be delimited to 48 bronchial asthma patients attending Lagos State University Teaching Hospital’s respiratory clinic, Ikeja, Lagos.

#### Significance of the study

It is expected that the outcome of this study will establish the relationship among Interferential therapy, bronchodilators, and body positions on Asthma Control, Asthma Quality of life, and selected pulmonary variables of patients living with bronchial Asthma. It is expected that this study will provide a piece of substantial evidence on airway reversibility in bronchial Asthma using IFT.

## METHOD

### Study Design

The study is a parallel, 12-week randomized control trial. The study will involve four (4) intervention groups and four parallel placebo control groups.

### Participants

The participants for this study will consist of male and female adult bronchial asthma patients aged 18 years and above attending Lagos State University Teaching Hospital (LASUTH), Ikeja, Lagos, Nigeria, West African.

*The inclusion criteria involve patients with bronchial Asthma aged 18 years and above attending the respiratory clinic of LASUTH*.

*The exclusion criteria involve patients with other types of COPD other than bronchial Asthma, hypersensitive to B2 agonist. Patients on a cardiac pacemaker, recent surgery, supplemental oxygen therapy, cardiac conditions, and patients with psychological impairments*.

Participants who met the required criteria will be asked to read and sign an informed consent approval for this study by the appropriate institutional review board.

### Setting

Patients with bronchial Asthma attending the respiratory clinic of Lagos State University Teaching Hospital Ikeja, Lagos State, Southwest Nigeria, would be recruited for the study. The hospital is a tertiary health facility within the state and receives referrals from within and outside the state.

### Sample size

Pulmonary function test is the primary outcome of interest for the study and the expected clinically relevant difference for pulmonary rehabilitation in various body positions using LLN and GLI reference equation proposed by Quanjer PH ^11^. Therefore, the sample size (N) will be determined using G-Power statistics software. The power is selected at 95% =0.95, confidence level at 5% =0.05 and effect size of 0.35 fig (1).

**Fig 1.**
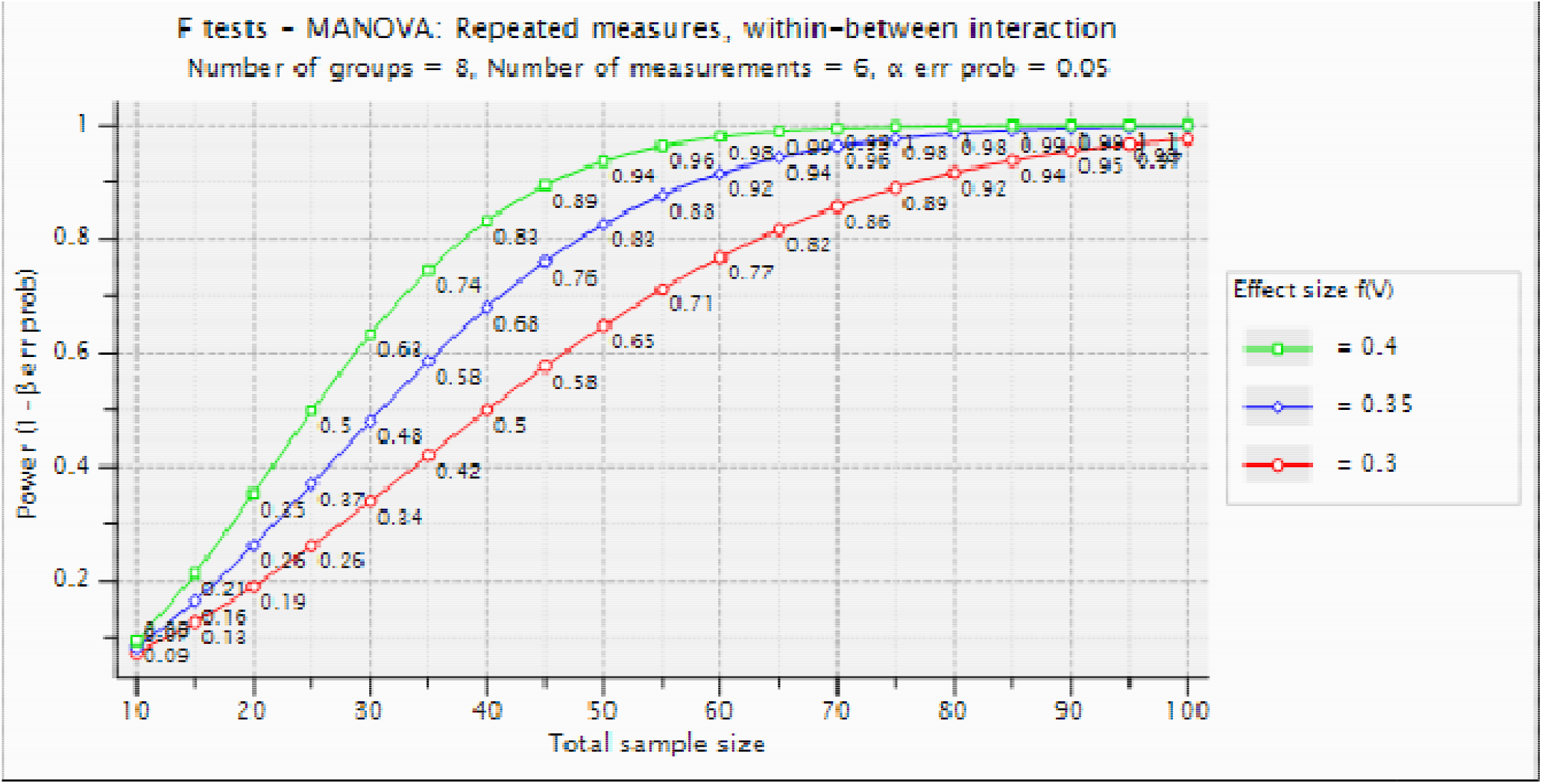
F tests -MANOVA: Repeated measures, within-between interaction.

### Randomization and blinding

The contact numbers of participants will be randomly extracted from the respiratory patients’ database attending Lagos State University Teaching Hospital Ikeja’s respiratory clinic (Figure 2). A bulk Text message captioned “Invitation to a study on ASTHMA will be circulated using the Luxury bulk SMS platform. Respondents will be assessed for eligibility, and those that meet the inclusion and exclusion criteria will participate in the study.

**Fig 2.**
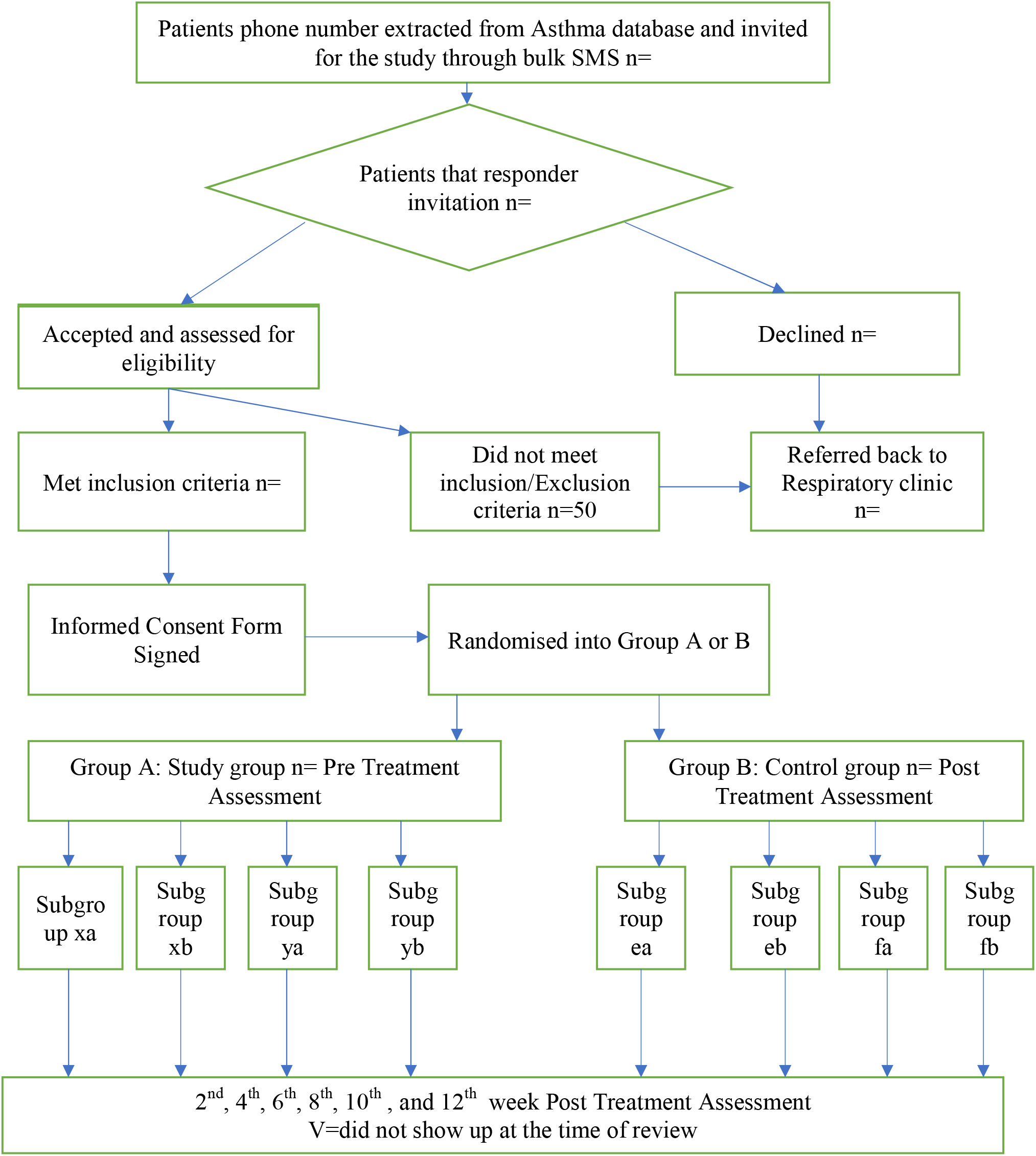
Recruitment and Randomization of Participants.

Participants will be randomly selected by simple randomization using a computer software program’s randomization table ^24^. The software program (www.randomization.com) will be used to allocate participants into study group A and control group B. Group A will be further assigned to subgroup x, y, z, and group B will be assigned to subgroup e, f, g. Subgroup x and e connote supine position, y and f connote 45°long sitting, while z and g represent 90° long sitting position, respectively.

### Procedure for Data collection

Forty-eight (48) subjects will be recruited for this study. The subjects will be randomly assigned into two major groups of twenty-four (24) subjects per group, two (2) subgroups of twelve (12) subjects per subgroup, and eight (8) final group of six (6) subjects per group.

### Assessment

The subjects’ medical records will be adequately screened for possible contraindications to the study. Subjects’ baseline respiratory parameters will be determined using the Koko PFT spirometer.

The Asthma Control Test questionnaire and Asthma quality of life will be administered to the control and study group at the beginning and every two weeks interval for 12 weeks of the study. The baseline spirometry will be conducted on the two groups before and after the commencement of the study.

Interferential unit “Nu-Tek E-Stim Pro” will be used for the study intervention. The treatment period will be increased by two minutes, with each application up to a total application time of 20 minutes.

### Assessment tools

#### Spirometry Assessment

The assessment will be conducted by a spirometrist certified by the Pan African Thoracic Society. A portable spirometer (Koko SX 1000 Standalone Version 7 Pneumotach) will be used to carry out this assessment. Daily calibration of the device will be conducted using a 3.0-liter syringe. A brief description of the assessment procedure, including technical steps to obtain pulmonary function data and variables, will be explained to each subject. After 2-3 tidal breaths, subjects will be asked to inhale deeply to total lung capacity and then exhale rapidly (without any pause) through a disposable mouthpiece until as much air as possible has been expelled from the lungs. The test will be performed in a sitting or standing position.

The assessments will be repeated three times after adequate rest. The maximum number of attempts permitted will be 8. After fulfilling the acceptability and repeatability criteria, we will select the two best curves. The average values of the forced vital capacity (FVC) and forced expiratory volume in the first second (FEV1) will be recorded ^25^.

#### Intervention

Participants will be briefed about the study’s nature, effect, and benefit of the study. They will be encouraged to clarify issues regarding the study. All participants will be required to give a written informed commence before participating in this study. Participants will be randomly assigned into two groups: Study (Group A) and Control group (Group B). Interferential therapy will be demonstrated to the study group alone. Asthma Control Test, Asthma Quality of life questionnaire, and Spirometry score will be measured and recorded before intervention in both groups. We will conduct a reassessment after the 2nd, 4th,6th, 8th, 10th, and 12th week of the study intervention.

The study group (Group A) will receive interferential therapy; this modality selection is only acceptable in the absence of cardiac disease history; it should not be used ^26^. In the absence of such a history, the subjects will be divided into two subgroups and labeled x and y. Subgroup ‘x’ will be allowed to use 400ug of SABA delivered in metered doses via a spacer 4-6 hours before the study, while subgroup ‘y’ will be allowed to use 24ug of LABA delivered in metered doses via a dry-powder inhaler within 24 hours of the study ^27^. In both cases, the inhaler technique, as described by ^28^ will be used to demonstrate the procedure to the participants. We will conduct the intervention with participants in a long sitting position, at an angle of 45° and 90° representing labels ‘a’ and ‘b,’ respectively. In both intervention positions, an electrode will be placed over the trapezius’s upper limits bilaterally on the upper back and the other two anteriorly over the lower ribs ^26^. The final group of participants to undergo the intervention will consist of ‘xa,’ ‘xb,’ ‘ya’ and ‘yb.’ If the subject is experiencing respiratory difficulty while the intervention is on-going, we will discontinue the procedure. With a 4,000 Hz base current, the interferential current range will be set from 10 to 150 Hz and initially applied for 10 minutes, carefully monitoring the patient’s condition during the treatment period. If the patient presents any sign of distress during treatment, we will turn the current off. As long as the subject experiences no distress with the IFC application, the treatment period will be increased by two minutes, with each application for up to 20 minutes.

The control group (Group B), in addition to the baseline pulmonary function test, will also receive counseling on Asthma and enjoy free musculoskeletal assessment. They will be divided into two subgroups and labeled ‘e’ and ‘f.’ subgroup ‘e’ will be allowed to use 400ug of SABA delivered in metered doses via a spacer 4-6 hours before the study, while subgroup ‘f’ will be allowed to use 24ug of LABA delivered in metered doses via a dry-powder inhaler within 24 hours of the study^27^. We will conduct the intervention with participants in a long sitting position, at an angle of 45° and 90° representing labels ‘a’ and ‘b,’ respectively. The final group of participants to undergo the intervention will consist of ‘ea,’ ‘eb,’ ‘fa’ and ‘fb.’ In both cases, the inhaler technique, as described by ^28^, will be used to demonstrate the procedure to the participants. They will be asked to maintain their respective positions for 20 minutes. If the subject is experiencing respiratory difficulty, we will discontinue the procedure.

#### Outcome Measures/Instruments

1. Asthma Control Test (ACT)
2. Asthma Quality of Life Questionnaire (AQLQ)
3. Spirometer (Koko SX 1000 Standalone Version 7 Pneumotach)
4. Interferential Therapy Machine (Nu-Tek E-Stim Pro)

#### 3.1.3 Description of Outcome Measures/Instruments

#### Asthma Control Test (ACT)

The asthma control test is a self-administered 5 item questionnaire developed for assessing asthma control level. It evaluates the most recent four-week time; each item is scored between 1 and 5, with a total score ranging from 5 to 25. An ACT score of 25 indicates that Asthma is “controlled,” whereas a score between 20 and 24 shows partially controlled Asthma, and a score of *<*20 indicates “uncontrolled” Asthma ^29^.

The ACT provides patients with Asthma, their doctors, and nurses with a useful measure to help determine the level of treatment required, it assesses the frequency of shortness of breath, night-time/early awakenings, rescue medication use, overall asthma control, and loss of productivity ^29, 30^. ACT has been clinically validated with spirometry and specialist assessment, it has been tested extensively in patients with Asthma and is recognized by the National Institutes of Health since its 2007 asthma guidelines ^29, 30^.

According to van van Dijk, Svedsater ^31^, there is a strong body of evidence supporting a relationship between improvement in ACT score and improved lung function, particularly with respect to forced expiratory volume in 1 s.

#### Asthma Quality of Life Questionnaire Standardised (AQLQ(S))

The Standardized version of the Asthma Quality of Life Questionnaire (AQLQS) is a 32-item questionnaire (self-administered or Clinician administered) with five domains, developed to measure the functional, Physical, Emotional, Occupational, and Social problems that are most troublesome to adults with Asthma ^18^. The maximum score obtainable in AQLQ(S) is 7.0, which translates to no impairment. The minimum score is 1.0, indicating severe impairment, 4.0 is the mid-range score, and indicates moderate impairment ^18^. We will utilize the Asthma Quality of Life Questionnaire’s standardized version to monitor the subjects’ problems in their daily lives.

## SPIROMETER

Koko SX 1000 Standalone Version 7 Pneumotach, a portable lightweight and comprehensive diagnostic tool, will be utilized to conduct a pulmonary function test. The Koko Legend II spirometer has a built-in thermal printer and a touch screen display; it can perform FVC, Pre vs. Post, and SVC tests. Test data and patient information are stored directly on an internal SD card that can be replaced and re-used. All the stored information can be downloaded via a USB cable onto a PC for backup or storage. This device supports daily calibration checks, complies with ATS-ERS 2005, has several Predicted Authors, and includes GLI-2012.

Daily calibration of the device will be conducted using a 3L syringe.

The participant’s condition can be shown by the ratio of the measured value and the predicted value. Flow rate-volume chart, volume-time chart display, data memory, delete, upload and review, trend chart display, scaling (Calibration), information prompts when volume or flow goes beyond the limits are features available in the device.

## BRONCHODILATORS

The administration of bronchodilators will be primarily through inhalation devices to deliver the drug to the lung’s bronchioles in metered doses. Inhalation devices come in all shapes and sizes, but critical is maximizing the amount of drug reaching the bronchioles. The best way to achieve maximum bioavailability is by fully exhaling, placing the inhaler in the mouth, and taking a full inhalation. After the patient has inhaled completely, it is followed by 10 seconds of no breathing to wait for the medicine to dissipate into the lung space. A slow exhalation back to normal breathing will be advised ^32, 33^.

### Inhaler technique

The eight-point technique listed below will be used to deliver bronchodilators and pre-BD reversibility testing in metered doses ^28^.

1. Remove the mouthpiece cover and shake
2. Hold the inhaler upright
3. Exhale to residual volume
4. Keep head upright or slightly tilted
5. Place mouthpiece between teeth and lips
6. Inhale slowly and press the canister
7. Continue slow and deep inhalation
8. Hold the breath for 5 seconds

According to Sim, Lee ^34^, albuterol (short-acting b2-bronchodilator) bronchodilator test, the subject will be required to exhale slowly fully and sprays an albuterol metered-dose of 100 μg (1 puff) while biting a valved chamber, after that, the subject will be required to slow and profoundly inhales until reaching maximum capacity (TLC) over 3–5 seconds, holds the breath for 5–10 seconds and exhales. The procedure will be repeated four times (total 400 μg of albuterol) at intervals of 30 seconds. However, if there is any concern about affecting the subject’s pulse rate or causing hand tremors, the dose may be decreased to 200 μg. After the last medication’s inhalation, the spirometry test is conducted again between 10–20 minutes ^35^.

## INTERFERENTIAL UNIT

The Nu-Tek E-Stim Pro provides low and medium frequency outputs from a single unit. Currents on the Nu-Tek E-Stim Pro include Interferential (2 and 4-pole), Russian, Diadynamic, TENS, Sinusoidal, Faradic, Galvanic, Interrupted Galvanic, Trabert, and Medi-Wave. The Solo Multidyne will be used to deliver interferential currents by generating a beat frequency range of 10-150hz from two medium frequency currents undulating at a base frequency of 4000Hz to 4100Hz ^14^. An interferential current will be activated with two electrodes placed posteriorly at the upper border of the trapezius and anteriorly below the ribs. A beat frequency will be generated at the point of intersection, resulting in relaxation of the smooth muscles, resolution of pain, and mobilization of secretions ^14^.

### Data analysis

We will use the Statistical Package for Social Sciences (SPSS Inc, Chicago, II) version 26.0 for the Windows package program to analyze data. Descriptive statistics of mean, standard deviation, frequency, and percentages will be used to summarize the results. Bar charts, pie charts, and histograms will be utilized for pictorial illustration. A multilevel analysis of variance (ANOVA) will be used to compare the outcome variables [body position (45 degrees long sitting and 90 degrees long sitting), pulmonary function variables (FEV1, FVC, FEV1/FVC), asthma control test (ACT), and standardized asthma quality of life questionnaire (AQLQ)] among each group, and dependent t-test will be used to compare the pre and post-test results while independent t-test will be used to compare the outcome variable across the two groups. We will set the level of significance at p0.05.

### Harms

This study carries minimal risks; the procedures are not life-threatening and should not cause harm or negative effect. This may include temporary muscle soreness, increased heart rate, blood pressure, sweating, and dizziness. Necessary care will be in place to prevent the occurrence of an adverse event. However, in case of a report of serious adverse events (e.g., comorbidities, injuries, persistent excruciating pain, dizzy spells, headache, etc.) after intervention or at any point during the trial, then we would consider unblinding the participant to the intervention for his/her safety. Additionally, the participants will be instructed to report any adverse events to the PI or the physiotherapist supervising their group. To ensure the participants’ adequate supervision and safety, we will limit the number of participants per group in a day to a maximum of 3. Arrangements have been made with the hospital’s Accident and Emergency units, where we will conduct the research to provide a standby medical team. However, the University of KwaZulu-Natal insurance scheme on clinical trials has fully covered the participants in this type of study.

## Discussion

The relationship between medication-induced airway reversibility and reversibility obtained from electrophysical modalities is still not well justified. Furthermore, the relationship between the mode of delivery of electrophysical agents and the recovery pattern in bronchial Asthma is yet to be fully understood.

A study by Karashurov, Gudovskii ^36^, programmed Electrostimulation of the sinocarotid nerves implanted to 78 patients with bacterial Asthma for six years was reported to have prevented the majority of asphyxia attacks, reduced their frequency by 2.7-fold, and the need for medications by 2.7-3.4-fold.

Aweto, Tella ^20^, in a study of IFT on cardiopulmonary parameters of 42 BA patients for six weeks reported a significant improvement in the systolic blood pressure (p=0.004), Forced Expiratory Volume in one second (p=0.02), Forced Vital Capacity (p=0.04), and Peak expiratory flow rate (p=0.007), while group B had significant reductions in these pulmonary parameters. There were significant improvements (increases) in the ACT score (p=0.0001) and AQLQ (p=0.001).

Mohammed and Elyazed ^21^, studied Thirty Egyptian children who have Asthma, and age ranged from 9-15 years, BMI 18.5 to 24.9 kilogram/ meter2. The pre- and post-treatment variables revealed a significant improvement in the three groups’ pulmonary functions with the favor of a laser puncture therapy group and interferential therapy group over diaphragmatic exercise.

Although studies by Aweto et al (2016), Karashurov, Gudovskii ^36^, and Mohammed and Elyazed ^21^ identified the effect of an electrophysical agent in the management of Asthma, their findings did not ascertain the possible effect of the medication used by the patients during the procedure. Consequently, it is expected that this study’s outcome will further reveal the effect of electrophysical modality on the symptoms frequently experienced by asthma patients who are on short or long-acting bronchodilator medication.

Finally, it is expected that the findings of this study will be a guideline for the management of BA with electrophysical agents and will further support the cost-benefit of Asthma management in Nigeria and other low-income countries.

## Data Availability

The corresponding author will make available the datasets for the study upon reasonable request. However, the findings from the study would be made available to participating researchers as required by law.

https://drive.google.com/drive/folders/12hZHwBlN0A27BcmAtxp1nGHHr2cVY_Jz?usp=sharing

## Access to Protocol

https://pactr.samrc.ac.za/TrialDisplay.aspx?TrialID=10942 The protocol was registered on 1st of May 2020 with identifier number PACTR202005807526130 and the trial organization is UKZN

## Ethical Considerations and consent to participates

The Biomedical Research Ethics Committee has approved this study of the University of KwaZulu Natal (South Africa) (Ethics Number: BREC/00001883/2020), and by the Human Research Ethics Committee of Lagos State University Teaching Hospital, Ikeja, Lagos, Nigeria, West Africa (LREC/06/10/1385). The study is registered with ClinicalTrial.gov with the following registration number: PACTR202005807526130.

A written and signed informed consent will be obtained from all participants recruited for this study through a third party that is independent of the study team. The consent form is designed by the Biomedical Research Ethics Committee of the University of KwaZulu-Natal (BREC) according to meet the WMA Helsinki declaration and good clinical practice (GCP). During this trial’s conduct, the PI will communicate in writing to the RECs in the event of the need to modify or amend the protocol, especially inclusion or exclusion criteria of the study.

## Consent to publication

Not Applicable

## Acknowledgments

We want to thank all the physiotherapists and medical doctors for their contribution to the study’s success and, most importantly, the participants’ voluntary participation. Our profound appreciation goes to Dr. Olufunke Adeyeye and Dr. Olufemi Ojo of Lagos State University Teaching Hospital, Ikeja, Lagos, for their professional advice in writing and reviewing this manuscript, also Miss Kemi and Miss Laide for their support in Pulmonary function test, and Miss Amodeni Ayomopewa for her editorial input.

## Abbreviations

ANOVA: Analysis of Variance
COPD: Chronic Obstructive Pulmonary Disease
FEV1: Forced expiratory volume in 1 second
FVC: Forced Vital Capacity
FRC: Functional residual Capacity
GLI: Global Lung Initiative
IFC: Interferential Current
IFT: Interferential Therapy
LASUTH: Lagos State University Teaching Hospital
LLN: Lower Level of Normal
MANOVA: Multiple Analyses of Variance
MS: Microsoft
PEFR: Peak expiratory flow rate
PFT: Pulmonary Function Test
RCT: Randomized Control Trial
ROM: Range of Motion
SPSS: Statistical Package for Social Sciences
UKZN: University of Kwazulu-Natal

